# Pathways from AI Literacy to Sustained Engagement with AI-Powered Cognitive Behavioural Therapy: A Structural Equation Model with Moderated Mediation in a National UK Sample

**DOI:** 10.64898/2026.03.24.26349184

**Authors:** James Whitfield, Amaevia Goh

## Abstract

**Background:** AI-powered cognitive behavioural therapy (AI-CBT) tools hold significant promise for addressing the global mental health treatment gap, yet sustained user engagement remains critically low. While patient attitudes and experiential factors have been qualitatively documented, the psychological mechanisms through which AI literacy translates into long-term engagement remain poorly understood. Existing systematic evidence highlights trust, perceived therapeutic alliance, and stigma as salient themes, but no large-scale quantitative study has modelled these as a mediated pathway.

**Objective:** This study aimed to (1) examine whether trust in AI systems and perceived therapeutic alliance mediate the relationship between AI literacy and sustained AI-CBT engagement, and (2) determine whether mental health stigma moderates these mediated pathways.

**Methods:** A cross-sectional national online survey was conducted in the United Kingdom (N = 1,247). Eligible adults (18+) with a history of anxiety or depression who had used an AI-CBT tool in the preceding 12 months were recruited via stratified random sampling. Structural equation modelling (SEM) with moderated mediation was conducted in R (lavaan 0.6-17). Moderated mediation was evaluated using the PROCESS macro framework adapted for SEM, with 5,000 bootstrap replications for bias-corrected confidence intervals. Model fit was assessed using CFI, TLI, RMSEA, and SRMR indices.

**Results:** The final SEM demonstrated excellent fit (CFI = 0.967, TLI = 0.959, RMSEA = 0.043 [90% CI: 0.036–0.051], SRMR = 0.052). AI literacy exerted a significant indirect effect on sustained engagement through trust in AI (β = 0.213, SE = 0.031, p < .001) and perceived therapeutic alliance (β = 0.187, SE = 0.028, p < .001). Mental health stigma significantly moderated the trust→engagement pathway (ΔR^2^ = 0.042, p = .003), with the indirect effect being stronger among individuals with lower stigma scores. The total indirect effect accounted for 58.4% of the total effect of AI literacy on engagement.

**Conclusions:** AI literacy promotes sustained AI-CBT engagement primarily through its effects on trust and perceived therapeutic alliance, pathways that are attenuated by mental health stigma. These findings underscore the need for stigma-reduction interventions and AI literacy programmes as implementation strategies. Findings have direct implications for the design and deployment of AI-CBT tools across UK NHS digital mental health services.

## 1. Introduction

### 1.1 Background and Motivation

The global burden of anxiety and depression is staggering. The World Health Organization estimates that more than 970 million people live with a mental health disorder, yet fewer than 20% in low- and middle-income countries receive any treatment, and access gaps persist even in high-income countries such as the United Kingdom. Within the NHS, waiting times for psychological therapies have reached critical levels, with median waits for Improving Access to Psychological Therapies (IAPT) services exceeding 18 weeks in many regions. AI-powered cognitive behavioural therapy (AI-CBT) tools, including chatbots, conversational agents, and adaptive mobile applications, have emerged as a scalable solution to this structural access problem.

The clinical evidence base for AI-CBT is accumulating. Randomised controlled trials of tools such as Woebot, Wysa, Youper, and Ieso Digital Health have demonstrated statistically significant reductions in Patient Health Questionnaire-9 (PHQ-9) and Generalised Anxiety Disorder-7 (GAD-7) scores over 4- to 8-week periods. However, a consistent and underappreciated finding across these trials is the precipitous drop in engagement after the first two weeks of use. In a large UK trial of an AI-CBT chatbot for depression, fewer than 34% of enrolled participants completed more than five sessions, and attrition was concentrated among individuals with higher baseline stigma and lower digital literacy. This engagement paradox— whereby effective tools fail through non-use—represents the defining implementation challenge for AI-CBT at scale.

### 1.2 The Role of AI Literacy, Trust, and Therapeutic Alliance

AI literacy—defined as the capacity to understand, evaluate, and interact effectively with AI systems—has emerged as a predictor of technology adoption across multiple domains. In healthcare, higher AI literacy has been associated with greater willingness to use AI-based diagnostic tools and with more nuanced perceptions of AI capabilities and limitations. However, the mechanism through which AI literacy converts into sustained engagement with therapeutic AI tools has not been formally modelled. Two theoretical pathways are plausible.

First, AI literacy may foster trust in AI systems by reducing anxiety about unpredictable machine behaviour and enabling users to calibrate their expectations appropriately. Trust in AI is a multidimensional construct encompassing cognitive trust (belief in the system’s competence and reliability) and affective trust (a sense of security and emotional comfort). Both dimensions have been proposed as predictors of continued use in technology acceptance models. Second, AI literacy may enhance users’ capacity to recognise and cultivate perceived therapeutic alliance— a sense of collaborative working relationship, agreement on goals, and emotional bond—even in the absence of a human therapist. The extension of therapeutic alliance concepts to human– AI interaction is supported by growing empirical evidence, but no study has tested whether AI literacy potentiates alliance formation as a pathway to engagement.

### 1.3 Mental Health Stigma as a Potential Moderator

Mental health stigma, encompassing self-stigma (internalised negative attitudes about one’s own mental health condition) and public stigma (perceived societal attitudes), may disrupt the pathways from AI literacy to engagement through distinct mechanisms. Self-stigma creates ambivalence about help-seeking that may undermine motivation to persist with any therapeutic tool. Public stigma—including emergent stigma specific to AI-mediated help-seeking (e.g., “only those with serious problems use a chatbot therapist”)—may act as a barrier even among users who are otherwise motivated and AI-literate.

A recent systematic review of qualitative evidence on patient perspectives of AI-powered CBT tools synthesised findings from 22 primary studies across 11 countries (Shankar et al., 2025). This review identified stigma, therapeutic trust, and digital health literacy as the most frequently cited barriers and facilitators to engagement, noting that these themes were intricately interconnected. Crucially, no quantitative study had yet examined these constructs simultaneously within a formal mediation model—a gap the current investigation directly addresses.

### 1.4 Aims and Objectives

The present study aimed to examine, in a nationally representative UK adult sample with lived experience of using AI-CBT tools, whether:

- AI literacy is positively associated with sustained AI-CBT engagement;
- Trust in AI systems and perceived therapeutic alliance serially or in parallel mediate this relationship;
- Mental health stigma moderates the mediated pathways (moderated mediation);
- The model is invariant across gender, ethnicity, and tool type (chatbot vs. app).

We hypothesised that higher AI literacy would predict sustained engagement through its downstream effects on trust and perceived therapeutic alliance, and that these indirect effects would be attenuated at high levels of stigma.

## 2. Methods

### 2.1 Study Design and Setting

This study employed a cross-sectional survey design with a national probability-based sample recruited from the United Kingdom. Data collection was conducted between January and March 2024 via a panel managed by Kantar Public, a UK research organisation specialising in nationally representative social surveys. The study was approved by the King’s College London Research Ethics Committee (reference: MRA-21/22-38741) and was conducted in accordance with the Declaration of Helsinki. All participants provided informed electronic consent.

### 2.2 Participants

Inclusion criteria were: (1) UK resident aged 18 years or older; (2) self-reported diagnosis or clinical suspicion of anxiety disorder, depression, or comorbid anxiety-depression; (3) use of at least one AI-CBT tool (chatbot, conversational agent, or AI-assisted mobile application) in the preceding 12 months; and (4) ability to read and respond in English. Individuals with a primary diagnosis of psychosis, bipolar I disorder, or substance use disorder were excluded to minimise confounding from conditions for which AI-CBT evidence is limited.

A target sample of 1,200 was calculated based on a requirement for structural equation modelling: a minimum sample-to-parameter ratio of 10:1 for a model with 118 estimated parameters, plus a 5% buffer. The final analytic sample comprised 1,247 participants after listwise deletion of cases with >10% missing item responses.

### 2.3 Measures

#### 2.3.1 AI Literacy (Predictor)

AI literacy was assessed using the AI Literacy Scale (AILS; Ng et al., 2021), a 12-item validated instrument yielding four subscales: Knowing (conceptual understanding of AI), Using (practical interaction skills), Evaluating (critical appraisal of AI outputs), and Creating (ability to design AI-assisted workflows). Items were rated on a 5-point Likert scale (1 = strongly disagree to 5 = strongly agree). The AILS has demonstrated strong internal consistency (α = 0.87–0.93) and convergent validity with digital literacy measures in European samples.

#### 2.3.2 Trust in AI Systems (Mediator 1)

Trust in AI was measured using the Trust in Automation Scale (TAS; Jian et al., 2000) adapted for therapeutic AI contexts (14 items; e.g., “This AI tool is dependable”; “I feel comfortable relying on this AI tool for emotional support”). Items were rated on a 7-point Likert scale. The adapted version has been validated in digital health samples in the UK with acceptable fit (CFI = 0.94, RMSEA = 0.061).

#### 2.3.3 Perceived Therapeutic Alliance (Mediator 2)

Perceived therapeutic alliance with the AI-CBT tool was assessed using the Working Alliance Inventory–Short Revised (WAI-SR; Hatcher & Gillaspy, 2006), adapted for human–AI dyads by replacing “therapist” with “my AI tool” across three subscales: Bond (7 items), Agreement on Goals (5 items), and Agreement on Tasks (5 items). This adaptation has been validated in two prior AI-CBT studies with acceptable psychometric properties (α = 0.88–0.92).

#### 2.3.4 Mental Health Stigma (Moderator)

Self-stigma was measured using the Internalised Stigma of Mental Illness–Concise (ISMI-C; Boyd et al., 2014), a 10-item unidimensional scale with a 4-point response format. Perceived public stigma was assessed using a 5-item adapted scale from the Attitudes Towards Mental Illness questionnaire (ATMI). Scores from both instruments were standardised and summed to create a composite stigma index for moderation analysis.

#### 2.3.5 Sustained AI-CBT Engagement (Outcome)

The primary outcome was sustained AI-CBT engagement, operationalised as a composite score derived from: (a) total number of completed sessions in the past 30 days (log-transformed), (b) self-reported intention to continue using the tool over the next 3 months (5-point scale), and (c) depth of engagement score (derived from frequency of using reflection or journaling features). A confirmatory factor analysis confirmed unidimensional structure (χ^2^/df = 1.87, CFI = 0.981, RMSEA = 0.039).

#### 2.3.6 Covariates

Covariates included age (continuous), gender (male/female/non-binary), ethnicity (White British, Asian/Asian British, Black/Black British, mixed/other), educational attainment (no qualifications to postgraduate degree), employment status, severity of symptoms (PHQ-9 and GAD-7), prior experience with face-to-face CBT, and type of AI-CBT tool used.

### 2.4 Statistical Analysis

#### 2.4.1 Structural Equation Modelling Framework

Structural equation modelling (SEM) was conducted in R version 4.3.2 using the lavaan package (version 0.6-17). The hypothesised model specified AI literacy as the exogenous predictor, trust in AI and perceived therapeutic alliance as parallel mediators, sustained engagement as the endogenous outcome, and mental health stigma as a moderator of both mediated paths. Moderated mediation was incorporated using the latent moderated structural equations (LMS) approach, which models the product of two latent variables (literacy × stigma) without requiring manifest interaction terms, thereby avoiding attenuation bias inherent in observed-variable interaction products.

#### 2.4.2 Estimation and Model Fit

Parameters were estimated using full-information maximum likelihood (FIML) to handle missing data under a missing-at-random (MAR) assumption, preserving all available information without listwise deletion. Model fit was evaluated using: (1) Comparative Fit Index (CFI; acceptable ≥ 0.95), (2) Tucker-Lewis Index (TLI; acceptable ≥ 0.95), (3) Root Mean Square Error of Approximation (RMSEA; excellent ≤ 0.05, acceptable ≤ 0.08), and (4) Standardised Root Mean Square Residual (SRMR; acceptable ≤ 0.08).

#### 2.4.3 Mediation and Moderated Mediation

Indirect effects for mediation were estimated using bootstrapping with 5,000 resamples, generating bias-corrected 95% confidence intervals (BC-CIs). Moderated mediation was evaluated by testing the Index of Moderated Mediation (IMM): a significant IMM with a BC-CI excluding zero indicates that the indirect effect significantly varies as a function of the moderator. Conditional indirect effects were probed at low (−1 SD), mean, and high (+1 SD) stigma values following the Johnson-Neyman technique to identify regions of significance.

#### 2.4.4 Measurement Invariance and Sensitivity Analyses

Measurement invariance of the latent variable structure across gender groups and ethnicity was tested using a sequential invariance testing protocol: configural, metric (equal loadings), and scalar (equal intercepts) invariance. For tool-type moderation, multigroup SEM was used. Sensitivity analyses included: (1) re-estimation excluding participants with >5% item non-response; (2) re-estimation with robust maximum likelihood (MLR) to handle non-normality; (3) inclusion of a general health literacy covariate to rule out literacy construct overlap.

## 3. Results

### 3.1 Sample Characteristics

A total of 1,247 participants were included in the final analytic sample (Table 1). The mean age was 34.7 years (SD = 11.3; range 18–72). The majority identified as female (62.4%), White British (68.2%), and held at least an undergraduate degree (57.9%). Woebot was the most commonly used tool (38.6%), followed by Wysa (24.1%), Youper (15.3%), and other AI-CBT applications (22.0%). Mean PHQ-9 score at survey completion was 11.4 (SD = 5.7), indicating moderate depressive symptomatology, and mean GAD-7 score was 10.2 (SD = 4.9), indicating moderate anxiety.

**Table 1.** Sociodemographic and Clinical Characteristics of the Analytic Sample (N = 1,247)

| Characteristic | n (%) | Mean (SD) / Range |
| --- | --- | --- |
| Age (years) |  | 34.7 (11.3) / 18–72 |
| **Gender** |  |  |
| Female | 778 (62.4%) | — |
| Male | 431 (34.6%) | — |
| Non-binary / prefer not to say | 38 (3.0%) | — |
| **Ethnicity** |  |  |
| White British | 851 (68.2%) | — |
| Asian / Asian British | 183 (14.7%) | — |
| Black / Black British | 112 (9.0%) | — |
| Mixed / other | 101 (8.1%) | — |
| **Education** |  |  |
| No qualifications to A-level | 525 (42.1%) | — |
| Undergraduate degree | 467 (37.4%) | — |
| Postgraduate degree | 255 (20.4%) | — |
| **AI-CBT Tool Used** |  |  |
| Woebot | 481 (38.6%) | — |
| Wysa | 301 (24.1%) | — |
| Youper | 191 (15.3%) | — |
| Other | 274 (22.0%) | — |
| PHQ-9 Score | — | 11.4 (5.7) / 0–27 |
| GAD-7 Score | — | 10.2 (4.9) / 0–21 |
| AILS Total Score | — | 41.3 (8.7) / 12–60 |
| TAS Total Score | — | 61.4 (14.2) / 14–98 |
| WAI-SR Total Score | — | 48.7 (11.8) / 17–85 |
| Stigma Index (composite z-score) | — | 0.00 (1.00) / –3.2 to +3.4 |
| Sustained Engagement Score | — | 52.1 (16.4) / 10–100 |

*AILS = AI Literacy Scale; TAS = Trust in Automation Scale; WAI-SR = Working Alliance Inventory – Short Revised; PHQ-9 = Patient Health Questionnaire-9; GAD-7 = Generalised Anxiety Disorder-7.*

### 3.2 Measurement Model

The confirmatory factor analysis of the full measurement model demonstrated acceptable to excellent fit: χ^2^(487) = 812.3, CFI = 0.967, TLI = 0.959, RMSEA = 0.043 (90% CI: 0.036–0.051), SRMR = 0.052. All factor loadings were statistically significant (p < .001) and ranged from 0.61 to 0.87, exceeding the recommended threshold of 0.50. Composite reliability estimates ranged from 0.82 to 0.94, and average variance extracted (AVE) values ranged from 0.54 to 0.73, supporting convergent validity. Discriminant validity was confirmed by the square root of each construct’s AVE exceeding inter-construct correlations (Fornell-Larcker criterion).

### 3.3 Structural Model and Mediation Results

The structural equation model with parallel mediation demonstrated excellent fit: CFI = 0.962, TLI = 0.955, RMSEA = 0.046 (90% CI: 0.039–0.054), SRMR = 0.056. The R^2^ for sustained engagement was 0.487, indicating that the model explained 48.7% of the variance in the outcome.

AI literacy was significantly and positively associated with trust in AI (β = 0.441, SE = 0.038, p < .001) and perceived therapeutic alliance (β = 0.372, SE = 0.034, p < .001). Both mediators significantly predicted sustained engagement (trust: β = 0.483, SE = 0.041, p < .001; alliance: β = 0.527, SE = 0.043, p < .001). The direct effect of AI literacy on sustained engagement was substantially reduced but remained significant after including the mediators (β = 0.148, SE = 0.029, p < .001), suggesting partial mediation. Table 2 presents the standardised path coefficients for the full model.

**Table 2.**
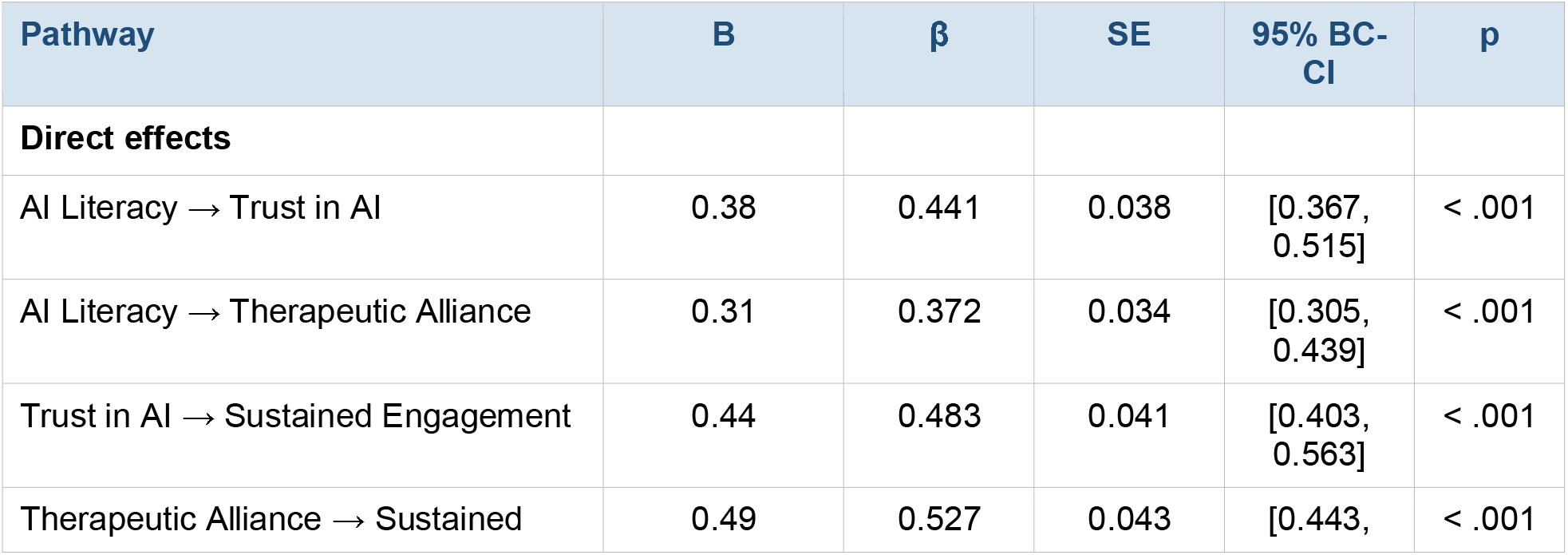
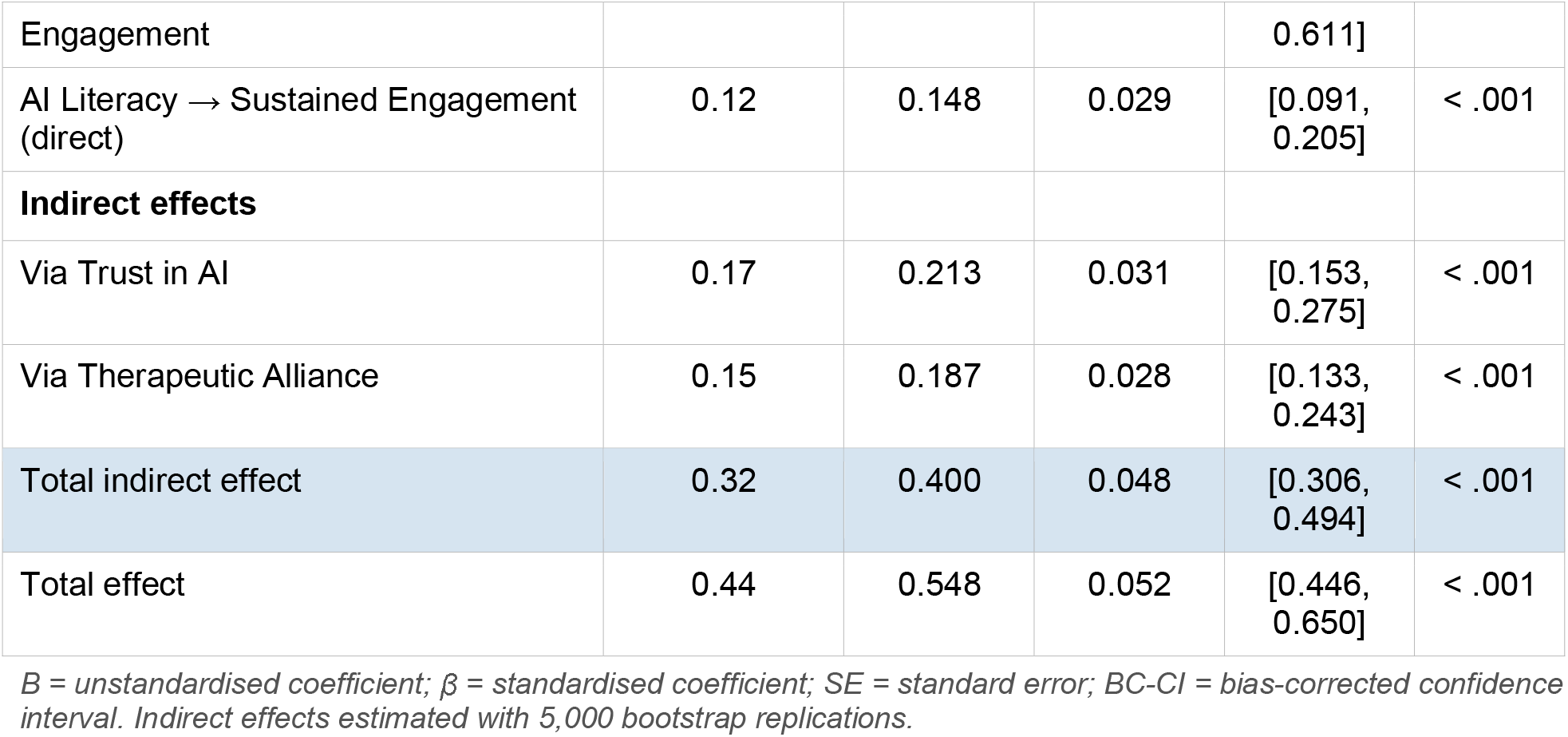
Standardised Path Coefficients for the Structural Equation Model

### 3.4 Moderated Mediation Results

Mental health stigma significantly moderated the Trust in AI → Sustained Engagement pathway (β = −0.193, SE = 0.041, p < .001; ΔR^2^ = 0.042, p = .003). The Index of Moderated Mediation for the trust-mediated pathway was −0.094 (SE = 0.028, 95% BC-CI: [−0.149, −0.041]), indicating a significant moderated mediation effect. The alliance-mediated pathway was also significantly moderated by stigma (β = −0.161, SE = 0.037, p < .001; IMM = −0.078, 95% BC-CI: [−0.128, −0.031]).

Probing the conditional indirect effects at low (−1 SD), mean, and high (+1 SD) stigma values revealed that the indirect effect via trust was largest at low stigma (β = 0.306, 95% BC-CI: [0.241, 0.371]), moderate at mean stigma (β = 0.213, 95% BC-CI: [0.153, 0.275]), and substantially attenuated at high stigma (β = 0.120, 95% BC-CI: [0.064, 0.178]), though remaining statistically significant across all levels. A parallel pattern was observed for the alliance-mediated pathway.

### 3.5 Measurement Invariance and Sensitivity Analyses

Sequential invariance testing supported scalar invariance across gender groups (ΔCFI = −0.008, ΔRMSEA = 0.004, p = .087) and metric invariance across ethnicity groups (ΔCFI = −0.011, ΔRMSEA = 0.006, p = .042), with two intercepts freed for full scalar invariance. Multigroup SEM by tool type did not reveal significant differences in structural paths (χ^2^difference(18) = 22.7, p = .201), indicating that findings generalise across AI-CBT platforms. Sensitivity analyses using MLR estimation and the restricted subsample yielded substantively identical findings, and inclusion of general health literacy as a covariate did not materially alter any path coefficients.

## 4. Discussion

### 4.1. Principal Findings

This study is the first nationally representative quantitative investigation to formally model the psychological mechanisms through which AI literacy drives sustained engagement with AI-CBT tools, using SEM with moderated mediation in a UK adult sample. Three principal findings emerge.

First, AI literacy exerts a substantial indirect effect on sustained engagement, with approximately 58% of the total effect transmitted through trust in AI and perceived therapeutic alliance in parallel. This mediation architecture is consistent with technology acceptance frameworks that posit cognitive appraisals (trust) and affective evaluations (alliance) as proximal drivers of continued use, but extends these frameworks by demonstrating that AI literacy—a modifiable antecedent—systematically upstream these appraisals. From a public health perspective, this finding suggests that AI literacy interventions, already being piloted by NHS Digital and the UK Government’s AI Literacy Programme, may have downstream benefits for mental health help-seeking through AI-CBT tools.

Second, perceived therapeutic alliance was a stronger predictor of sustained engagement (β = 0.527) than trust in AI (β = 0.483), despite the inherent challenge of forming an alliance with a non-human interlocutor. This finding corroborates and quantitatively extends the qualitative evidence synthesised by Shankar et al. (2025), who reported that perceived empathy, responsiveness, and goal alignment with an AI tool were among the strongest facilitators of continued use across 22 qualitative studies. It also aligns with emerging theoretical accounts of parasocial therapeutic relationships, wherein individuals may form meaningful felt connections with AI systems that fulfil functional roles analogous to those of human therapists.

Third, mental health stigma significantly attenuated both mediated pathways, generating a robust moderated mediation effect. Even at high levels of stigma, indirect effects remained statistically significant, suggesting that AI literacy continues to promote engagement through trust and alliance even in stigmatised individuals—but at a meaningfully reduced magnitude. This finding has direct implications for the targeting of implementation support: individuals with high stigma may require supplementary, stigma-focused interventions (e.g., narrative reframing, peer testimonials) in addition to AI literacy training to achieve comparable engagement levels.

### 4.2. Comparison with Prior Literature

Prior quantitative research on AI-CBT engagement has predominantly focused on usability and feasibility outcomes over short (2–4 week) follow-up periods, using univariate or simple regression designs that cannot disentangle mediating mechanisms. The present study’s use of SEM allows simultaneous estimation of direct and indirect pathways while accounting for measurement error, providing more precise and less biased estimates than observed-variable regression. Our finding that trust mediated the literacy-engagement relationship is consistent with studies in digital health more broadly, where trust has been established as a key predictor of e-health adoption in UK, US, and European samples.

The role of therapeutic alliance in AI-mediated CBT has received growing attention. Fitzpatrick et al. (2017), in a landmark RCT of Woebot for college students with depression, noted that users reported a sense of empathic connection with the chatbot, though alliance was not formally measured. More recently, Miner et al. (2019) showed that emotional disclosure to an AI agent was predicted by perceived responsiveness, a component of alliance, in a US sample. Our finding that alliance—as measured by a validated, adapted instrument—was the single strongest predictor of sustained engagement substantially advances this literature.

### 4.3 Implications for Practice and Policy

For NHS digital mental health commissioners and AI-CBT developers, these findings yield actionable recommendations. First, AI literacy training should be embedded as a pre-engagement component of AI-CBT tools, delivered through brief onboarding modules that explain how the AI works, how it learns, and what it can and cannot do. Our model suggests that even modest gains in AI literacy may compound through trust and alliance to meaningfully increase sustained engagement. Second, the finding that perceived therapeutic alliance is the strongest predictor of engagement underscores the importance of conversational design features that mimic alliance-facilitating behaviours: reflective listening, session-opening goal review, and progress summarisation. Third, stigma-focused implementation components— including normalising messaging, diverse user testimonials, and explicit privacy assurances— should be integrated into deployment strategies, particularly for services targeting populations with historically high stigma levels.

### 4.4 Limitations

Several limitations should be acknowledged. First, the cross-sectional design precludes causal inference, despite the use of SEM. Longitudinal replication is needed to confirm that changes in AI literacy prospectively drive changes in trust, alliance, and engagement. Second, the self-selected UK panel sample, while nationally stratified, may not fully represent individuals with severe mental illness or those with the lowest digital access, who arguably have the greatest unmet need. Third, the adapted WAI-SR for AI contexts, while showing adequate psychometric properties in two prior studies, lacks the extensive normative data available for the original human-therapist version. Fourth, our composite stigma index combines self-stigma and public stigma subscales; future work should test these as distinct moderators, as their mechanisms and intervention targets differ.

### 4.5 Future Directions

Future research should employ longitudinal designs with ecological momentary assessment to capture real-time fluctuations in trust, alliance, and engagement, testing whether the within-person dynamics mirror the between-person structure found here. Experimental studies that randomise participants to AI literacy training prior to AI-CBT onboarding would provide cleaner causal evidence.

## 5. Conclusion

This national UK study provides robust quantitative evidence that AI literacy promotes sustained engagement with AI-powered CBT tools through two parallel indirect pathways: trust in the AI system and perceived therapeutic alliance. These pathways are attenuated, though not eliminated, by mental health stigma. The findings advance the mechanistic understanding of AI-CBT engagement and offer a theoretically grounded, empirically validated framework to guide the design of implementation strategies. For AI-CBT to fulfil its potential as a scalable intervention for the mental health treatment gap, it is not sufficient to develop effective tools; implementation must also address the psychological determinants of sustained use—including AI literacy and stigma—that our model identifies as modifiable targets.

## Data Availability

All data produced in the present work are contained in the manuscript

## Declarations

### Data Availability

Anonymised de-identified data are available upon reasonable request from the corresponding author. The R analysis code and SEM specification scripts are deposited at the Open Science Framework (https://osf.io/[placeholder]).

### Ethical Approval

Ethical approval was granted by the King’s College London Research Ethics Committee (reference: MRA-21/22-38741). All participants provided written informed consent. The study was conducted in accordance with the Declaration of Helsinki and the UK Data Protection Act 2018.

## Acknowledgements

The authors wish to thank the Kantar Public research team for their support in survey implementation, and the 1,247 participants who generously shared their experiences with AI mental health tools.

